# Importance of patient bed pathways and length of stay differences in predicting COVID-19 bed occupancy in England

**DOI:** 10.1101/2021.01.14.21249791

**Authors:** Quentin J. Leclerc, Naomi M. Fuller, Ruth H. Keogh, Karla Diaz-Ordaz, Richard Sekula, Malcolm G Semple, ISARIC4C Investigators, CMMID COVID-19 Working Group, Katherine E. Atkins, Simon R. Procter, Gwenan M. Knight

## Abstract

**Objectives:** Predicting bed occupancy for hospitalised patients with COVID-19 requires understanding of length of stay (LoS) in particular bed types. LoS can vary depending on the patient’s “bed pathway” - the sequence of transfers between bed types during a hospital stay. In this study, we characterise these pathways, and their impact on predicted hospital bed occupancy.

**Design:** We obtained data from University College Hospital (UCH) and the ISARIC4C COVID-19 Clinical Information Network (CO-CIN) on hospitalised patients with COVID-19 who required care in general ward or critical care (CC) beds to determine possible bed pathways and LoS. We developed a discrete-time model to examine the implications of using either bed pathways or only average LoS by bed type to forecast bed occupancy. We compared model-predicted bed occupancy to publicly available bed occupancy data on COVID-19 in England between March and August 2020.

**Results:** In both the UCH and CO-CIN datasets, 82% of hospitalised patients with COVID-19 only received care in general ward beds. We identified four other bed pathways, present in both datasets: “Ward, CC, Ward”, “Ward, CC”, “CC” and “CC, Ward”. Mean LoS varied by bed type, pathway, and dataset, between 1.78 and 13.53 days.

For UCH, we found that using bed pathways improved the accuracy of bed occupancy predictions, while only using an average LoS for each bed type underestimated true bed occupancy. However, using the CO-CIN LoS dataset we were not able to replicate past data on bed occupancy in England, suggesting regional LoS heterogeneities.

**Conclusions:** We identified five bed pathways, with substantial variation in LoS by bed type, pathway, and geography. This might be caused by local differences in patient characteristics, clinical care strategies, or resource availability, and suggests that national LoS averages may not be appropriate for local forecasts of bed occupancy for COVID-19.

## Introduction

Patients with Coronavirus disease 2019 (COVID-19) can display a range of symptoms which vary in severity (1), with up to one in five infections requiring hospitalisation, and up to 16% of hospitalised patients requiring intensive care (2,3). However, hospital bed capacity is limited; for example, there was only approximately 1 critical care bed per 10,000 inhabitants in England as of February 2020 (4). If demand for beds exceeds hospital capacity, this would severely decrease the quality of healthcare provided, and lead to worse outcomes for all patients who require hospitalisation, not only those with COVID-19. Consequently, it is essential to be able to accurately predict demand for hospital beds, both to allow a pre-emptive scale up of capacity and to act as a barometer for the effectiveness of upstream public health measures (5,6).

To prioritise resource allocation, models that can project bed occupancy are used at the hospital, regional and national level (7–12). For COVID-19, these models typically use two simplifying assumptions; first, that a patient’s hospital stay occurs in one bed type (e.g. general ward bed or critical care bed), and second, that the stay of each patient lasts for a fixed period. However, these assumptions are now inconsistent with publicly available data showing that many patients occupy different bed types for varying durations of stay (13).

Here, we use both hospital and national level COVID-19 inpatient data to identify the range of “bed pathways”, the sequence of transfers between bed types, for a single hospital stay. We then estimate the length of stay (LoS) by bed type for each stage in these pathways. Finally, we develop and apply a model to examine the importance of using these bed pathways for bed occupancy predictions, notably for key indicators such as peak bed requirements and capacity required over time.

## Methods

All of the analyses below were conducted in R (14), using the following packages: openxlsx (15), here (16), dplyr (17), reshape2 (18), linelist (19), ggplot2 (20), cowplot (21), knitr (22), rlist (23) and mstate (24). The code for the bed occupancy model and bed pathways data are available in a GitHub repository (https://github.com/qleclerc/COVID_bed_occupancy).

### 1. Patient bed pathways

A bed pathway is the sequence of transfers between bed types during a hospital stay. We considered two bed types used in acute care hospitals in England: general ward and critical care (CC) beds. Note that in England, CC beds include both intensive care unit (ICU) beds and high dependency unit (HDU) beds (13), but in other countries “CC beds” and “ICU beds” can be equivalent terms (25). An example of a pathway in our analysis would be “Ward, CC”, where a patient is admitted first to a ward bed, then to a CC bed, before being discharged or dying.

#### UCH

We derived patient bed pathways from two datasets. Our first dataset was provided by University College Hospital (UCH), a teaching hospital that is part of the University College London Hospitals NHS Foundation Trust, and one of the designated hospitals in London for admission of patients with COVID-19. This first dataset contains date of admission, type of bed used and data of discharge information on 168 inpatients with COVID-19 admitted to UCH between 6th March 2020 and 17th April 2020. All of these patients are now discharged or dead, therefore the LoS from this dataset were uncensored. The patients are not identifiable in this dataset, and are only associated with a number between 1 and 168. If a patient stayed in more than one bed type during their hospitalisation, the dataset includes an admission date, bed type, and discharge date for each stage.

To identify all possible bed pathways, we gathered all recorded stages by patient, and ordered them in chronological order. For each patient, the LoS for each stage was then estimated by taking the difference between the admission date and discharge date and adding one (such that a patient admitted and discharged on the same day would have an LoS of 1). The data was regrouped by bed pathway, and for each pathway we calculated the number of stages and the proportion of total patients following the pathway. Finally, for each stage in each bed pathway we calculated the mean LoS and standard deviation (s.d.) To compare these estimates with those typically used in bed occupancy models (7–12), we also calculated an “average LoS by bed type”, which is the mean LoS of all stages of that bed type recorded across all pathways. For sensitivity, we also estimated median LoS and interquartile range for each stage and pathway.

#### CO-CIN

The second dataset was from the ISARIC WHO CCP-UK study (National Institute for Health Research Clinical Research Network Central Portfolio Management System ID: 14152) an ongoing prospective cohort study in England, Scotland, Wales and Northern Ireland (13). The dataset we used contains information from 208 acute care hospitals. We used a subset of this data, restricted to patients in the COVID-19 Clinical Information Network (CO-CIN). These were patients with proven or high likelihood of SARS-CoV-2 infection admitted to hospital between 11th March 2020 (the date when hospital admission policy in England changed from admitting all known COVID-19 infected to “when necessary”) and 19th July 2020: a total of 42,980 individuals. This dataset covers 80% of all patients with COVID-19 hospitalised in England during this period.

For the CO-CIN dataset, multi-state modelling analyses were used to estimate the time patients spent in each bed type, before transitioning to a different type, as well as to discharge or in-hospital death (26,27). Using this, we estimate LoS for each stage in each possible bed pathway. These models account for competing risks and censoring, as not all patients had complete follow-up, notably because some patients were still hospitalised at the end of the data collection. The number of stages in each bed pathway, and the number of patients following each pathway were also recorded.

This resulted in a set of empiric LoS distributions, one for each stage in each bed pathway. We drew 100,000 samples from each empiric distribution to estimate mean LoS and s.d. for each stage. As with the dataset from UCH, we again calculated an “average LoS by bed type”, which is the mean LoS of all stages of that bed type recorded across all pathways. For sensitivity, we also estimated median LoS and interquartile range for each stage and pathway.

#### International

To obtain an international comparison of the possible types of bed pathways and the proportions of patients going through each, we used studies collated in a systematic literature review of LoS values for patients with COVID-19 (28). From these studies, mostly from China, we attempted to extract possible pathways and patient proportions (see Supplementary Material).

### 2. Model outline

To examine the impact of using patient bed pathways on bed occupancy forecasts, we developed a flexible discrete-time model. This model takes as input a time series of the number and date of observed (or forecast) daily hospital admissions over any given period of time, and, for each bed pathway *i*, the following characteristics:

– the probability that any patient will go down this pathway (*p*_*i*_, where ∑_*i*_ *p*_*i*_ = 1)
– the number of stages for each pathway *i* (*N*_*i*_)
– an average LoS in each stage *j* for each pathway *i* (*d*_*i,j*_)
– a standard deviation for each LoS (*s*_*i,j*_)

For example, the pathway “Ward, CC” contains two stages (N_W,CC_ = 2) with a first stage where the patient stays in a ward bed for an average of *d*_{W,CC},1_ days and s.d. of *s*_{W,CC},1_ days, followed by a second stage in a CC bed for an average of *d*_{W,CC},2_ days, s.d. *s*_{W,CC},2_.

The model then proceeds as follows: on the first day (*t*_0_) of the admission data time series, we assign a pathway, *k*, to each admitted patient by sampling from all of the possible pathways, weighted by the probabilities, *p*. Then, for each patient, we randomly assign a LoS for each of the *N*_*k*_ stages by sampling from a Weibull distribution (commonly used to represent LoS distributions (28)) with mean *d*_*k,j*_ (with *j* in *N*_*k*_) and standard deviation *s*_*k,j*_, and record that a bed of the corresponding type will be needed for that duration and for the specific time window determined by the order of the stages. Note that if a bed is needed for a fraction of a day, we conservatively assume that it is needed for that entire day (effectively rounding up the LoS to the next day). For sensitivity, we explored the impact of using a Lognormal instead of a Weibull distribution, and of rounding the LoS to the nearest day instead of rounding up.

Alternatively, instead of the mean and S.D., we can specify an empirical distribution for the LoS in each stage of the bed pathway (which we use with CO-CIN bed pathways data - see below). In that case, for each pathway *i* and stage *j*, the model takes as input a vector of values for the distribution (*a*_*i,j*_) and a vector of the corresponding probabilities for each value in the distribution (*r*_*i,j*_). For a patient assigned to a pathway, *k*, the LoS at each stage, *j*, is then sampled from the values provided in *a*_*kj*_, weighted by the probabilities provided in *r*_*kj*_.

For all results, 100 simulations of the model are performed, for which we report the daily mean overall bed requirements and 95% confidence intervals (mean +/-1.96 * standard error). For each bed type, we also calculate across these 100 simulations the median time and mean size of peak bed occupancy during the period over which admissions are generated, alongside the interquartile range (IQR) and standard deviation for peak time and peak size respectively.

### 3. Bed occupancy predictions

#### UCH

To obtain a time-series of daily hospital admissions and bed occupancy for UCH between 6th March 2020 to 17th April 2020, we re-used the same UCH dataset on COVID-19 inpatients described above. To generate the admissions time-series, we counted the earliest recorded admission date for each patient as one hospital admission on that date, regardless of bed type and bed pathway. Our time-series of admissions therefore represents admission to hospital, rather than admissions to a specific bed type. To generate a time-series of general ward and CC bed occupancy, we looked at each bed pathway recorded for each patient. For each stage in each pathway, we noted the hospital bed type that was occupied by the patient during that stage (general ward or CC), and recorded that one hospital bed of that type was occupied between the beginning and end dates for that stage.

We then compared the time-series of UCH bed occupancy to model-predicted occupancy for the same period. This serves both as a model validation step, and an opportunity to explore the impact of patient bed pathways as a service evaluation for which model to use for UCH.

Model-predicted occupancy for UCH was generated using the UCH hospital admissions time-series, and either the UCH bed pathways characteristics, or the UCH average LoS by bed type values. The UCH bed pathways characteristics used to parameterise the model (number of pathways, number of stages for each pathway, bed type at each stage, and mean LoS and standard deviation for each stage) were derived from our analysis above (see “Methods - Patient bed pathways”). This also applies to the average LoS by bed type estimates.

As a sensitivity analysis, we repeated this, keeping the UCH hospital admissions time-series, but using CO-CIN bed pathways characteristics and average LoS by bed type, instead of the UCH equivalents.

#### CO-CIN

As CO-CIN covers patients across the UK, we use the LoS estimates to predict bed occupancy at a national level. To support this analysis, we used publicly available data from the UK Government COVID-19 dashboard on daily hospital admissions in England (total, not separated by bed type) between 19th March 2020 and 19th August 2020 (29). This dashboard also provides data on bed occupancy by bed type, from 20th March 2020 to 19th August 2020 for total occupancy (general ward plus CC), and from 2nd April 2020 to 19th August 2020 for CC occupancy only (29). We derive the number of patients in general ward beds by subtracting the CC occupancy from the total occupancy.

We then compared the time-series of England bed occupancy to model-predicted occupancy for the same period. Model-predicted occupancy for England was generated using the England hospital admissions time-series, and either the CO-CIN bed pathways characteristics, or the CO-CIN average LoS by bed type values. The CO-CIN bed pathways characteristics were derived from our analysis above (see “Methods - Patient bed pathways”), and provide empirical distributions from which to sample LoS at each stage in each pathway. On the other hand, the CO-CIN average LoS by bed type values only rely on mean LoS and standard deviation.

As a sensitivity analysis, we repeated this, keeping the England hospital admissions time-series, but using UCH bed pathways characteristics and average LoS by bed type, instead of the CO-CIN equivalents.

#### Regional analysis

The UK Government COVID-19 dashboard also provides daily hospital admissions and bed occupancy for each of the seven NHS Regions in England, and for 208 NHS Trusts (29,30). We repeated the above analysis for each NHS Region, using either the CO-CIN bed pathways characteristics, or average LoS by bed type values. As this highlighted potential heterogeneity in LoS between Regions (see “Results”), we then searched for the best-fitting LoS values for each Region. We used the average LoS by bed type values, and allowed these to vary between NHS Regions, while maintaining the original CO-CIN proportions of patients going into either a general ward or CC bed. We identified the best-fitting values for LoS by minimising the sum of squared differences between model-predicted bed occupancy and publicly available bed occupancy data by bed type, calculated for each day available. Similarly, we found Trust level variation and hence searched for the best-fitting LoS values for each NHS Trust, to determine within-NHS Region heterogeneity.

As a sensitivity analysis, we also fit the model-predicted bed occupancy at the NHS Regions level by adjusting the proportion of patients going to a CC bed in each Region, while maintaining the original CO-CIN average LoS by bed type values.

## Results

### 1. Patient bed pathways

#### UCH

We identified five possible bed pathways for the 168 patients with COVID-19 admitted to UCH between 6th March 2020 and 11th April 2020 (Table 1). Most patients followed the “Ward” pathway (n = 137, 81.5%), only staying in a ward bed during their hospitalisation at UCH. The other patient pathways, ranked from most to least common, were: “Ward, CC, Ward” (6.0%), “Ward, CC” (5.3%), “CC” (4.8%) and “CC, Ward” (2.4%).

**Table 1:**
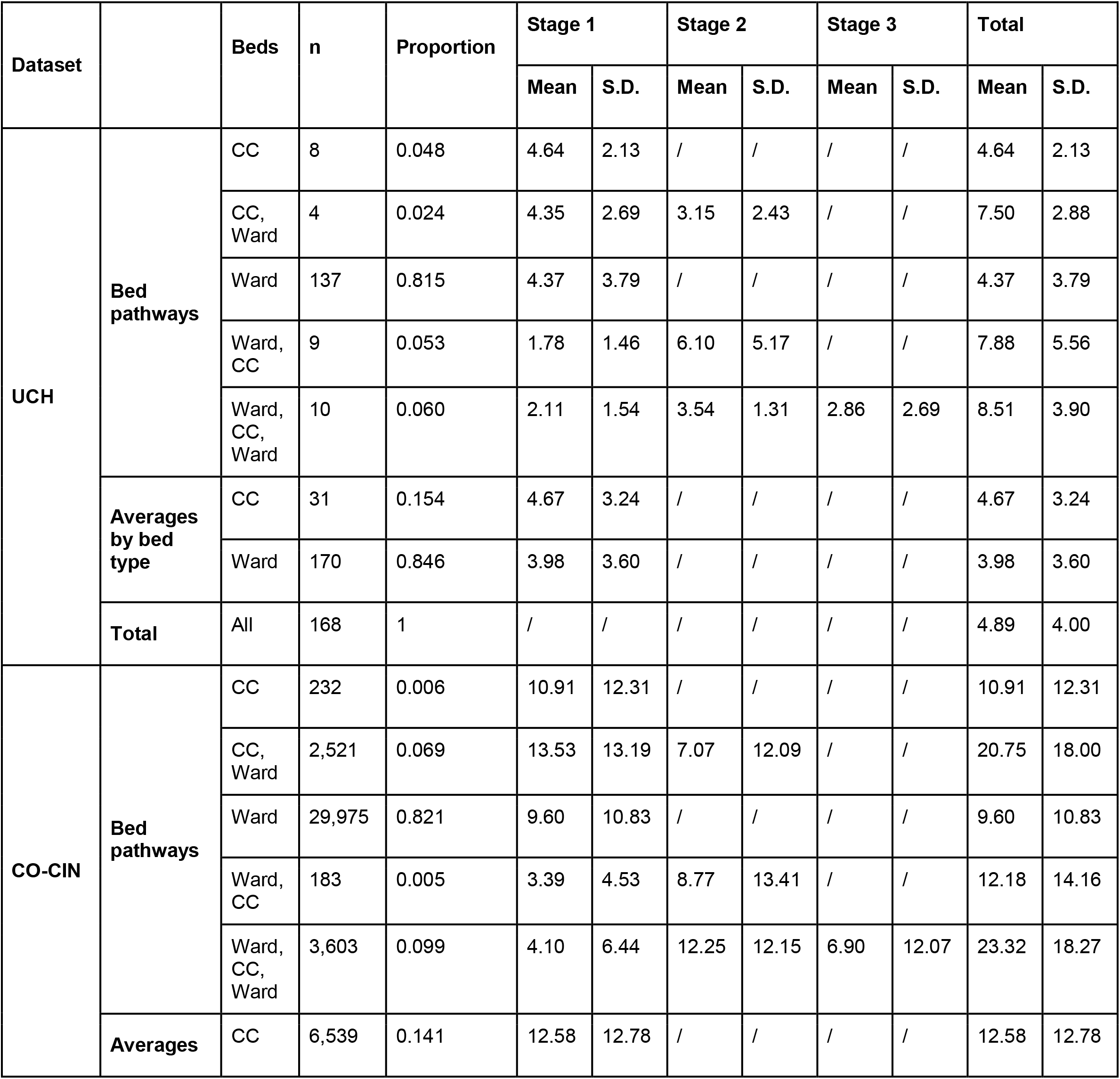
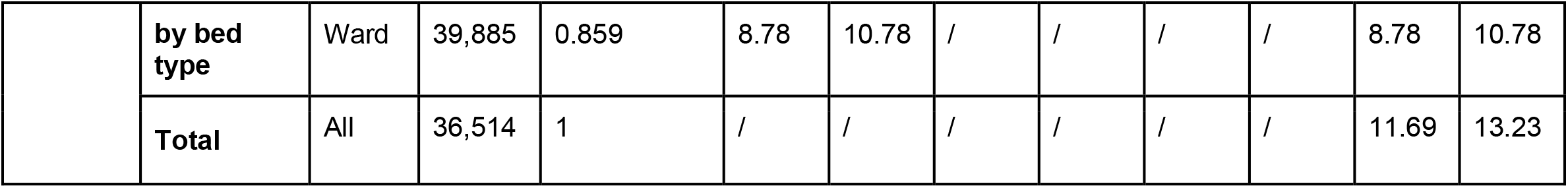
Patient pathways and length of stay for patients with COVID-19 from University College Hospital (UCH) and the COVID-19 Clinical Information Network (CO-CIN). CC: critical care. n: number of occurrences of that pathway (for Bed pathways), or bed type (for Averages by bed type). Note that the sum of n for the bed pathways differs from the sum for the averages, since two stages of the same bed type in one pathway correspond to two occurrences of that bed type in the averages. S.D.: standard deviation.

The mean LoS in a ward bed across all pathways and all stages was 3.98 days (standard deviation (s.d.) 3.60 days), while the mean LoS in a CC bed was 4.67 days (s.d. 3.24). However, LoS in each bed type varied between different pathways (Table 1 and Figure 1). The shortest mean length of stay in a ward bed was recorded in the “Ward, CC” pathway (1.78 days, s.d. 1.46), and the longest was recorded in the “Ward” pathway (4.37 days, s.d. 3.79). For CC beds, the shortest mean LoS was recorded in the “Ward, CC, Ward” pathway (3.54 days, s.d. 1.31), and the longest was recorded in the “Ward, CC” pathway (6.10 days, s.d. 5.17). Median LoS and IQR values are presented in Supplementary Table 2.

**Table 2:** Peak bed occupancy predicted using bed pathways data for patients with COVID-19 from UCH and CO-CIN. Daily hospital admissions are taken from UCH or UK Government data at the England level. Peak is defined as the earliest time at which maximum bed occupancy is reached. Results are from 100 model runs. Date format is dd/mm/yy. CC: critical care. occ.: occupancy. S.D.: standard deviation.

| Dataset | LoS used | Ward peak bed occupancy |  |  |  | CC peak bed occupancy |  |  |  |
| --- | --- | --- | --- | --- | --- | --- | --- | --- | --- |
|  |  | Median peak time | IQR peak time | Mean peak occ. | S.D. peak occ. | Median peak time | IQR peak time | Mean peak occ. | S.D. peak occ. |
| UCH | Bed pathways | 03/04/20 | 31/03/20 - 03/04/20 | 54 | 4.06 | 02/04/20 | 31/03/20 - 04/04/20 | 13 | 2.51 |
|  | Averages | 31/03/20 | 30/03/20 - 03/04/20 | 46 | 3.51 | 31/03/20 | 30/04/20 - 03/04/20 | 11 | 2.12 |
|  | Data | 31/03/20 | N/A | 59 | N/A | 31/03/20 | N/A | 16 | N/A |
| CO-CIN | Bed pathways | 10/04/20 | 09/04/20 - 10/04/20 | 20,469 | 89.89 | 14/04/20 | 13/04/20 - 15/04/20 | 4,530 | 65.05 |
|  | Averages | 10/04/20 | 10/04/20 - 10/04/20 | 18,209 | 99.07 | 11/04/20 | 10/04/20 - 12/04/20 | 3,835 | 56.26 |
|  | Data | 12/04/20 | N/A | 16,093 | N/A | 12/04/20 | N/A | 2,881 | N/A |

**Figure 1:**
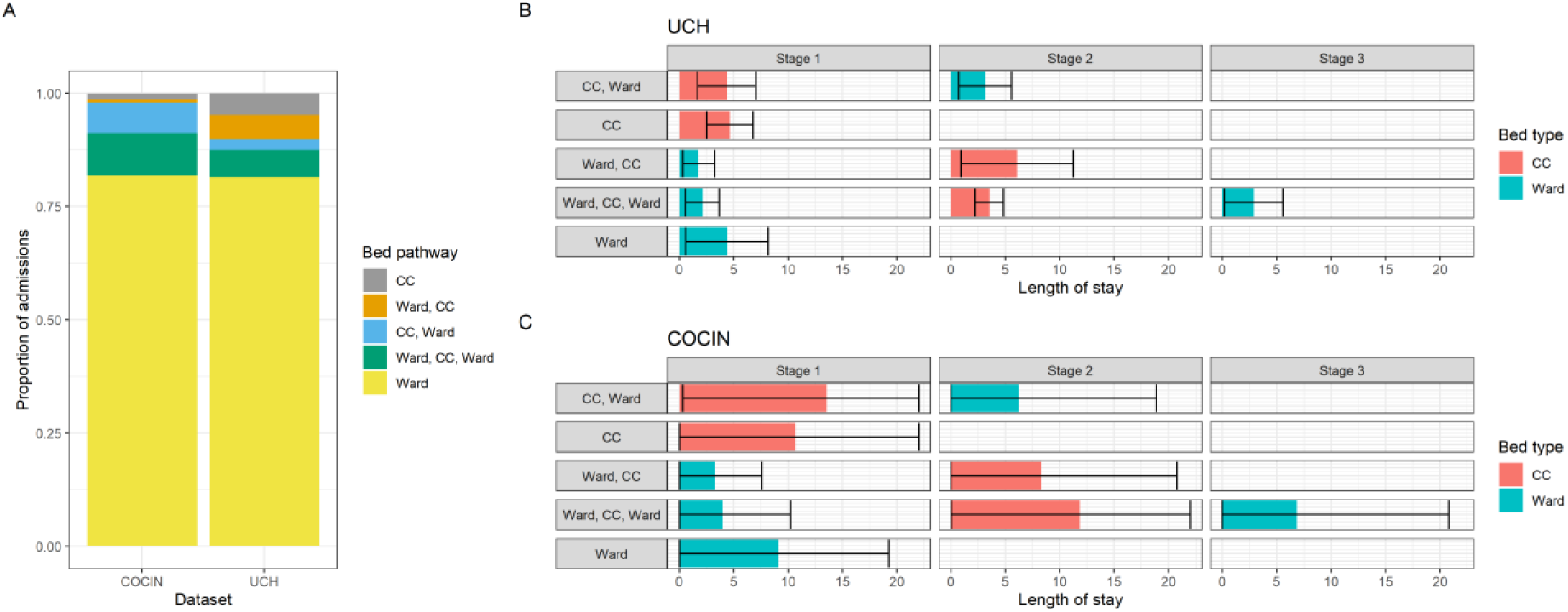
The proportions of admissions entering each bed pathway are similar for UCH and CO-CIN, but CO-CIN has longer length of stays by bed type and pathway. **A)** The proportion of admissions entering each bed pathway for UCH and CO-CIN. **B)** & **C)** Length of stay by stage in pathway (columns) and bed type (colour) for patients with COVID-19 from University College Hospital (UCH, **B**) and the COVID-19 Clinical Information Network (CO-CIN, **C**). The five distinct pathways are detailed in the grey boxes on the left. Error bars indicate mean plus or minus standard deviation, and are capped at 0 and 22.

#### CO-CIN

We found the same five bed pathways using the CO-CIN dataset (Table 1). Most patients followed the “Ward” pathway (n = 29,975, 82.1%); however, the proportion of patients going through the other bed pathways differ slightly compared to UCH: “Ward, CC, Ward” (9.9%), “CC, Ward” (6.9%), “CC” (0.6%) and “Ward, CC” (0.5%).

LoS estimates and standard deviations are higher for all stages and all bed pathways in CO-CIN compared to UCH (Table 1 and Figure 1). Across pathways, the mean LoS in a ward bed was 8.36 days (s.d. 10.53), and 12.31 days in a CC bed (s.d. 12.47). Again, we see variation in LoS by bed type between pathways (Figure 1). The shortest mean length of stay in a ward bed was recorded in the “Ward, CC” pathway (3.27 days, s.d. 4.32), and the longest was recorded in the “Ward” pathway (9.08 days, s.d. 10.19). For CC beds, the shortest mean LoS was recorded in the “Ward, CC” pathway (8.28 days, s.d. 12.49), and the longest was recorded in the “CC, Ward” pathway (13.53 days, s.d. 13.22). The complete empirical and multistate-modelled distributions for these LoS obtained from the CO-CIN dataset can be seen in Supplementary Figure 1. Median LoS and IQR values are presented in Supplementary Table 2.

#### International

In our analysis of studies reporting length of stay, none gave bed pathway information but 21 did give information on the “proportion of patients ever requiring an ICU bed” (see Supplementary Material). The weighted mean by patient number over these was 14% (s.d. 8%) (Supplementary Table 1). This is consistent with the aggregate “proportions of patients ever requiring a CC bed” we found in UCH (18.5%) and CO-CIN (17.9%).

### 2. Impact of patient bed pathways on predicted bed occupancy

#### UCH

Model-predicted bed occupancy for UCH is similar to, but underestimates, true occupancy (Figure 2 A-B). Using bed pathways, compared to average LoS by bed type, resulted in values closer to the data (Figure 2 A-B and Supplementary Table 3). In addition, predicted peak bed demand values were higher, and hence more accurate, by approximately 17% and 18%, for ward and CC beds respectively, when using bed pathways compared to average LoS by bed type (Table 2).

**Table 3:** Best-fitting length of stay values by NHS Regions. The England weighted average is the average of fitted LoS values in NHS Regions weighted by the proportion of cumulative England hospitalisations that occurred in each Region. CC: critical care.

| Source of LoS values | Geography | Ward bed LoS (mean) | CC bed LoS (mean) |
| --- | --- | --- | --- |
| CO-CIN | *England average* | 8.78 | 12.58 |
| Best fit by Region | East of England | 7.55 | 9.18 |
|  | London | 6.97 | 12.64 |
|  | Midlands | 7.36 | 8.08 |
|  | North East and Yorkshire | 7.58 | 8.54 |
|  | North West | 8.57 | 8.09 |
|  | South East | 7.87 | 8.07 |
|  | South West | 7.85 | 9.22 |
|  | *England weighted average* | 7.62 | 9.31 |

**Figure 2:**
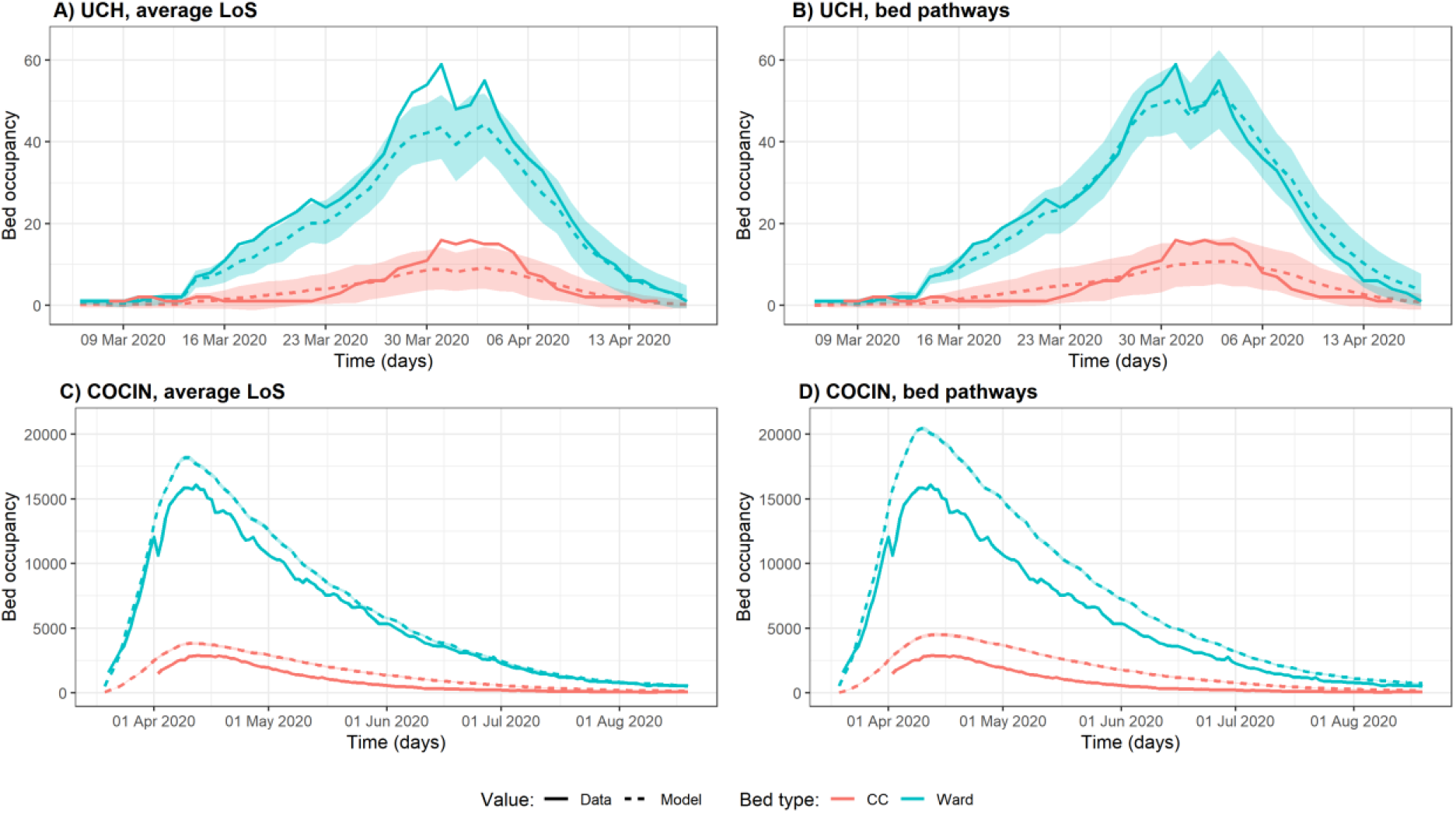
Using bed pathways instead of average length of stay by bed type affects model-predicted bed occupancy. Bed occupancy at UCH and model-predicted bed occupancy using UCH **A)** average length of stay estimates or **B)** bed pathways. Bed occupancy in England and model-predicted bed occupancy using CO-CIN **C)** average length of stay estimates or **D)** bed pathways. Shaded area is the 95% confidence interval from 100 model runs. Note that the time period is different between data from UCH and England.

For ward beds, 95% of data points lie within the 95% confidence interval of the model-predicted bed occupancy using bed pathways, while for CC beds this value is 89%. The median peak time in ward bed occupancy estimated by the model is 3rd April 2020 (IQR 31st March - 3rd April) and 31st March 2020 (IQR 30th March - 3rd April), when using bed pathways or average LoS respectively, while the true peak occurred on 31st March 2020 (Table 2). For the CC bed occupancy peak, these dates were 2nd April 2020 (IQR 31st March - 4th April) and 31st March 2020 (IQR 30th March - 3rd April), when using bed pathways or average LoS respectively, and 31st March 2020 according to the data (Table 2).

#### CO-CIN

On the other hand, model-predicted bed occupancy for England using average LoS values from CO-CIN clearly overestimated ward and CC bed occupancy, compared to publicly available England hospitalisation data (Figure 2 C-D). Using bed pathways from CO-CIN instead of average LoS by bed type again led to higher estimates of bed occupancy, by approximately 12% and 18%, for ward and CC beds respectively (Table 2). The median estimated peak time for ward bed occupancy was 10th April 2020 when using either average LoS by bed type or bed pathways, compared to 12th April in the data. The number of beds needed at that time was estimated to be approximately 27% higher than the data when using bed pathways, or 13% when using average LoS estimates. As for peak CC bed occupancy, the data indicated this occurred on 12th April 2020, while the median peak times predicted by the model were 14th April (IQR 13th April - 15th April) and 11th April (IQR 10th April - 12th April), when using bed pathways or average LoS respectively. Mean peak CC bed occupancy was approximately 57% and 33% higher compared to the data when using bed pathways or average LoS respectively.

### 3. Heterogeneity in length of stay when predicting bed occupancy at a regional level

To further investigate discrepancies between model-predicted bed occupancy and data in England, we apply the model at the NHS Regions level using the values from CO-CIN. As at the national level, our model consistently overestimates ward and CC bed occupancy across all Regions, when using either average LoS or bed pathways, although the extent of this overestimate varies between NHS Regions (Figure 3 & Supplementary Figure 2).

**Figure 3:**
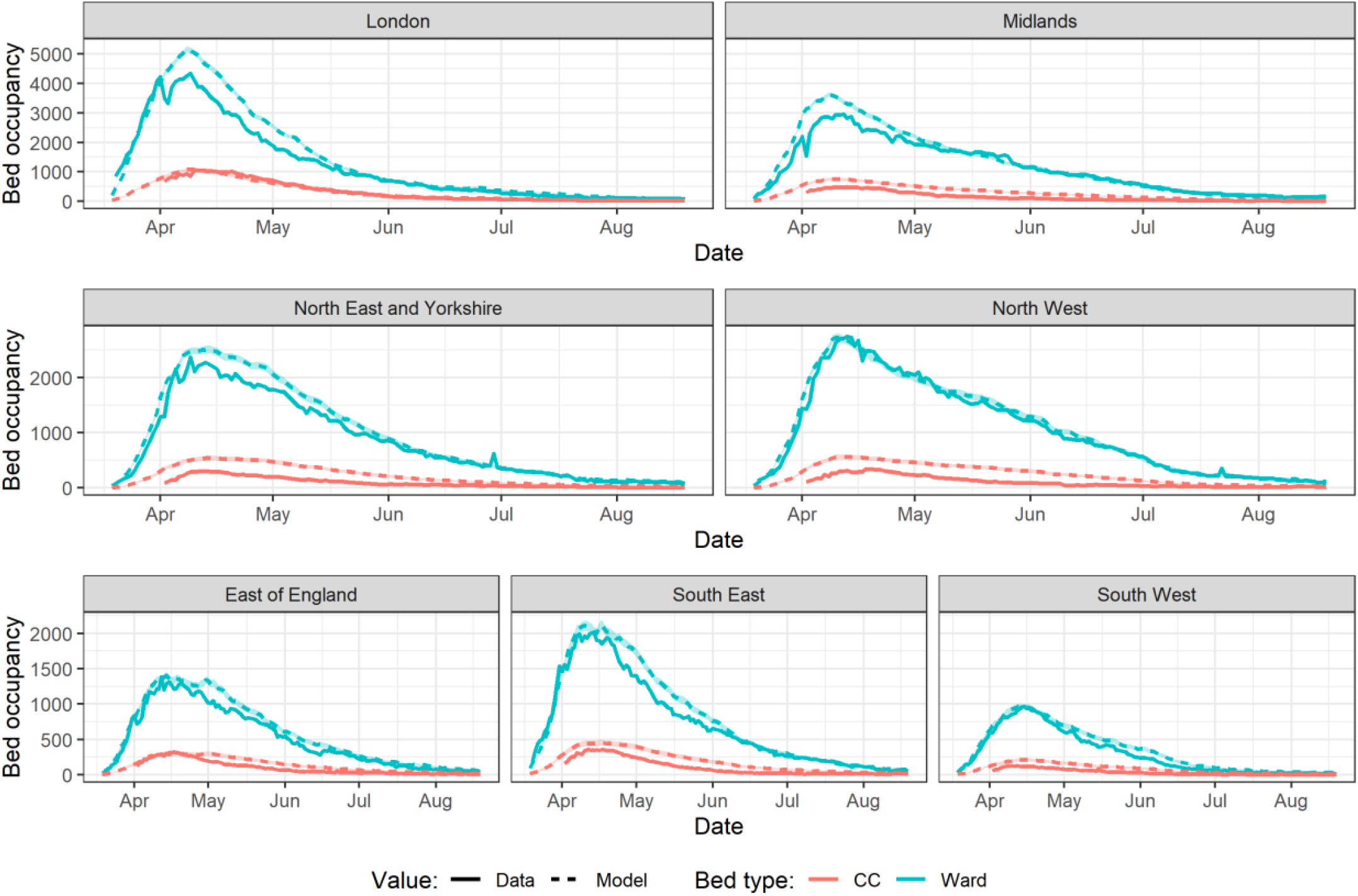
Model-predicted bed occupancy at the NHS Region level using average LoS values from CO-CIN. Full line shows true bed occupancy for the same period according to publicly available hospitalisation data (https://coronavirus.data.gov.uk/), with the dashed line being the mean of model output and 95% confidence interval (shaded area) from 100 runs. CC: critical care. Results are from 100 model runs. Results using bed pathways are qualitatively similar (Supplementary Figure 2).

Generating model estimates for average LoS for both bed types by fixing proportions of patients admitted to each bed type to those in CO-CIN and fitting bed occupancy for each Region separately brings the model-predicted bed occupancy much closer to the data (Figure 4, Supplementary Table 4). The resulting best-fit LoS values for ward beds are smaller in all Regions compared to the default CO-CIN value (though still higher than UCH values) (Table 3). Similarly, best-fit LoS values are smaller for CC beds in all Regions compared to the CO-CIN value, except for London. Recalculating averages for England using these regional LoS estimates weighted by the number of hospitalisations in each Region gives a similar pattern: LoS of 7.62 days for ward beds and 9.31 days for CC beds, while CO-CIN values for these are respectively 8.78 days and 12.58 days.

**Figure 4:**
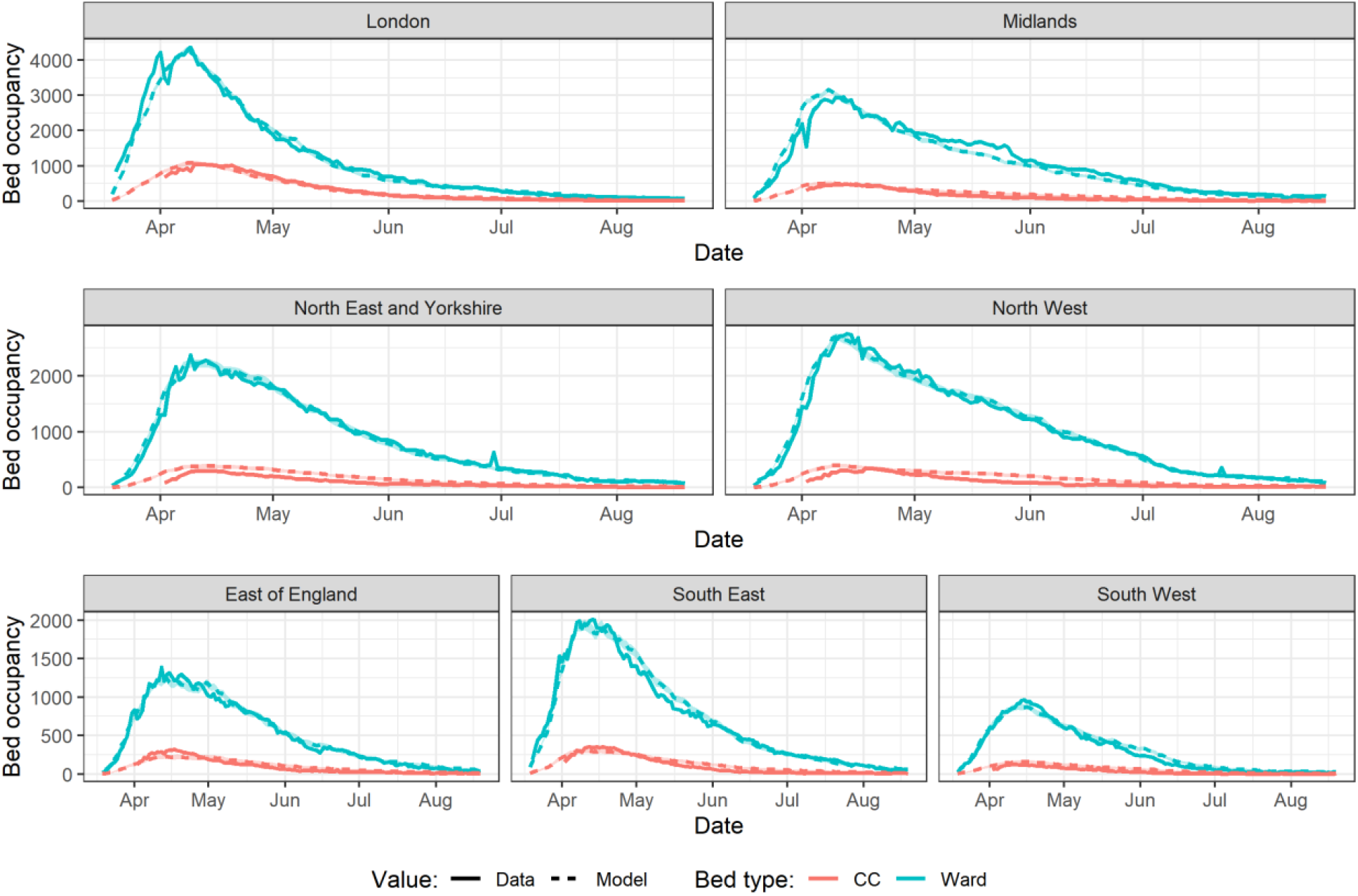
Model-predicted bed occupancy at the NHS Region level using average best-fit LoS values. Best-fit average LoS values were obtained by minimising the sum of squared differences between model values and data, for each bed type and Region separately. CC: critical care. Model outputs are mean and 95% confidence intervals (shaded region) from 100 model runs.

Figure 5 shows the distribution of best-fitting LoS values for 208 NHS Trusts, grouped by NHS Region. These boxplots indicate variations in LoS between NHS Trusts, and therefore within NHS Regions. The best-fitting LoS values for all the Trusts are available in the associated GitHub repository: https://github.com/qleclerc/COVID_bed_occupancy/blob/master/outputs/tables/best_fit_LoS_nhstrusts.csv.

**Figure 5:**
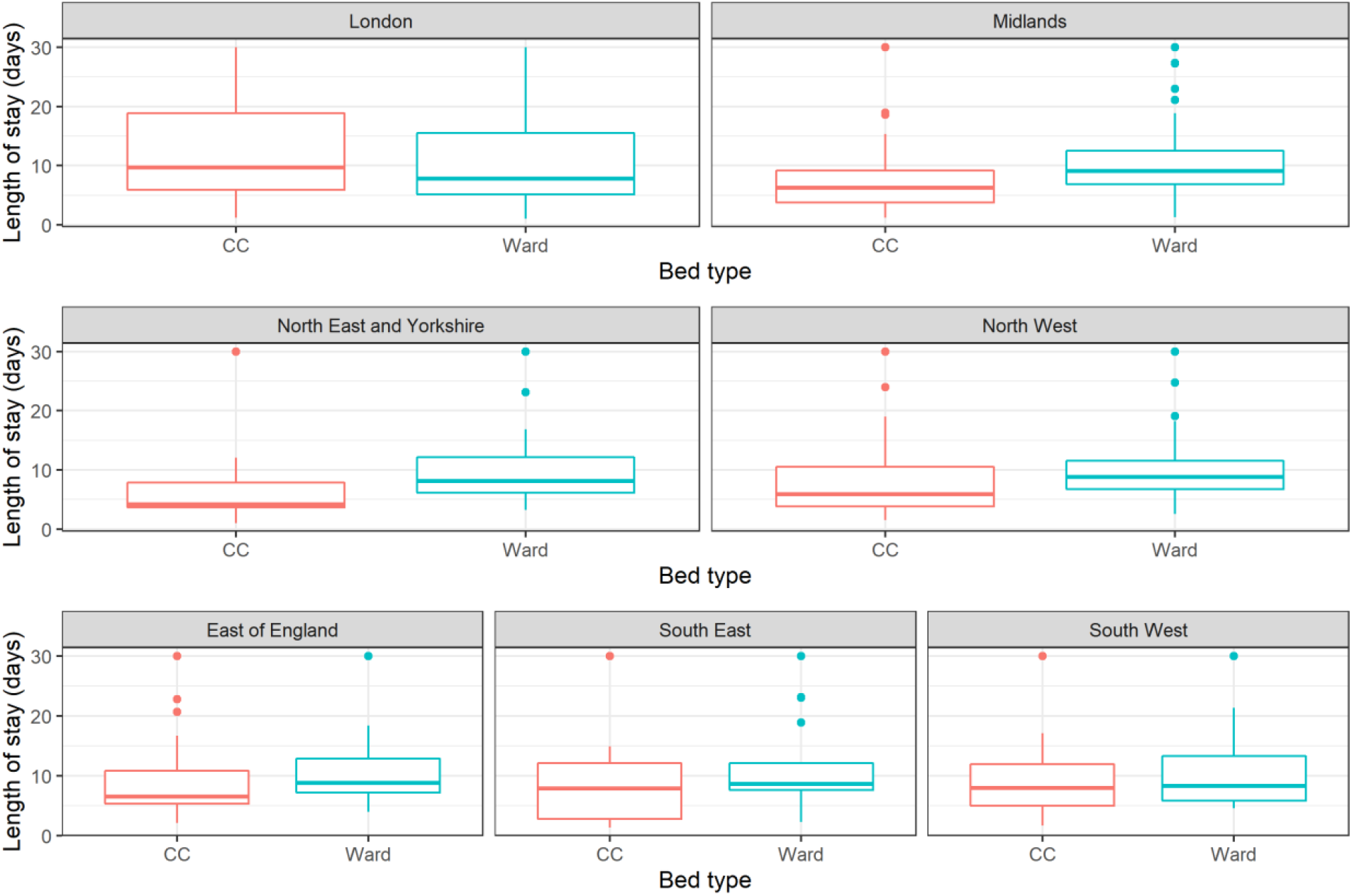
Distribution of best-fitting length of stay values by NHS Trusts, grouped by NHS Region. Best-fit average LoS values were obtained by minimising the sum of squared differences between model values and data, for each bed type and Trust separately. For each panel, the boxes correspond to the interquartile range (IQR) and the lines to a maximum of 1.5 times the IQR from the box limits. The centre lines in the boxes are the median. Maximum LoS is capped at 30 days. CC: critical care.

### 4. Sensitivity analysis

Our bed occupancy predictions were less accurate when using a Lognormal instead of a Weibull distribution for LoS sampling (Supplementary Figure 3, Supplementary Table 3).

We note that using UCH LoS estimates instead of CO-CIN to predict England bed occupancy does not improve the results, as this leads to an underestimate in bed occupancy (Supplementary Figure 4). Similarly, using CO-CIN LoS estimates to predict UCH bed occupancy leads to an overestimate in ward bed occupancy, and an underestimate in CC bed occupancy (Supplementary Figure 4).

Varying the proportion of patients requiring CC beds instead of the LoS estimates led to a worse fit of bed occupancy, although the resulting proportions were consistent with the data (Supplementary Figure 5, Supplementary Table 4, Supplementary Table 5).

Discretizing our LoS samples by rounding to the nearest day instead of rounding up to the next day (i.e. ceiling) naturally led to reduced bed occupancy predictions. In the case of UCH, ceiling instead of rounding improved the match to data (Supplementary Table 3).

## Discussion

### 1. Key findings

Accurate predictions of bed capacity, staff and associated equipment requirements in hospitals require an understanding of patient bed pathways and length of hospital stay. In this work we determined that, between March and August 2020 in England, most patients with COVID-19 (82%) required only a ward bed, for an average of 9.08 days. The remaining patients followed one of four other bed pathways, with lengths of stay (LoS) that varied by pathway and bed type. Incorporating bed pathways improved model-predicted bed occupancy at a local hospital level, compared to only using average LoS values by bed type. However, analysis at the England, NHS Regions and Trust level indicate that simply using national LoS values, even taking into account the bed pathways, did not generate reported regional or national bed occupancy.

### 2. Limitations

Our results are subject to limitations, notably linked to our model assumptions. Firstly, we chose to round-up sampled LoS estimates to the next day instead of rounding to the closest. In the case of UCH, this improved the fit of the model-predicted bed occupancy to the data (Supplementary Table 3), however in other settings this assumption could lead to overestimating bed occupancy. Nevertheless, we consider this to be a “safer” approach, as it implies that a bed needed for a fraction of a day is needed for that entire day. For capacity planning, slightly overestimating bed occupancy will likely be preferred to underestimating it, which could have severe consequences on clinical care. Secondly, we did not include a maximum bed capacity - this could improve accuracy if reaching limits frequently prevents patients from moving between bed types.

Thirdly, we assume that behaviours remain constant through time, which means that the average LoS estimates and proportion of patients going through each bed pathway do not change with time. This may not be realistic, as hospital discharge rates could vary with time, depending on capacity and changes in organisation. Similarly, the proportions of patients requiring critical care could change over time, depending on the public health measures in place which may affect the demographics of people most likely to be infected.

Finally, we assume independence within bed pathways, meaning that the LoS for each stage is drawn randomly, regardless of the LoS in previous stages. In reality, there could be a correlation between stages, where for example patients who stay in a CC bed for longer then need a longer recovery time in a ward bed, or vice versa.

On the data side, we are limited to making conclusions linked to the period of data collection: many things may now have changed for patients with COVID-19 such as differences in discharge, clinical care and admission status, and they may have changed during data collection. For a single hospital, estimating this time variance with small numbers is difficult, and we would expect stochasticity to have a greater impact, which would also explain why the model and data do not perfectly align for UCH. Nosocomial cases could also affect length of stay estimates and bed occupancy. However, we were interested in total bed occupancy for COVID-19 patients and hence did not exclude them from our data.

### 3. Results in context

We currently have limited published evidence on the length of hospital stay for patients with COVID-19, and even less on patient bed pathways: two reviews estimate the median LoS in a CC bed for patients with COVID-19 to be approximately 8 days (28,31) and median total hospital LoS to be 14 days (IQR 10-19) in China (28). Our estimates were shorter for UCH, but similar in CO-CIN. This could be due to right-censoring in the UCH dataset (only included those patients no longer in the hospital) or due to the fact that few studies with similar conditions and healthcare settings (i.e. outside China) were included in the above mentioned reviews. For comparison, average LoS in a CC bed for patients needing advanced respiratory support in England during 2018-19 is 4.9 days (32). The conclusion that most patients had care only in a general ward appears robust to international comparisons.

In the early months of the pandemic, the simplifying assumptions made by many capacity planning tools were necessary, as detailed data were not available on bed pathways. The next generation of bed capacity predictions should use patient bed pathways with local information on length of stay. To the best of our knowledge, there are only three other published models available which used patient bed pathways (33–35). The first one used the pathways “Ward”, “Ward, ICU (not ventilated)”, “Ward, ICU (ventilated)”, “Ward, ICU (not ventilated), Ward”, “Ward, ICU (ventilated), Ward” (33). However, the sources of the parameter values for these pathways are unclear, and seem to partly rely on a consensus of clinical experts, therefore comparisons with our results are difficult. On the other hand, the advantage of our analysis is that our code and length of stay data are both publicly available (https://github.com/qleclerc/COVID_bed_occupancy). As for the second model, it focused on machine-learning methods to estimate transition probabilities between the clinical states moderate/severe and critical, and is hence not comparable (34). The third model is closely aligned to our own work here, as it uses multi-state methods to estimate length of stay using a local hospital and a national dataset from the UK (35). Notably, this study investigates length of stay variations depending on when a patient was admitted to ICU during their hospitalisation, and also concludes that national estimates can differ from local ones.

### 4. Possible explanations and implications

We found that our initial hypothesis, that the LoS distribution for patients with COVID-19 should be the same everywhere in England, was unlikely to hold true. Instead, we observed regional variation, of up to 2 days differences for ward LoS and 4 days for CC LoS in the best fitting model estimates, as well as variation by NHS Trusts, and therefore within-NHS Regions. This could be explained in several ways. LoS for other diseases has been shown to vary between countries (36), and between hospitals, even after accounting for differences in local risk factors and demography (37).

However, the regional differences could also be due to true differences between NHS Regions in the COVID-19 care pathway and clinical practice. They could also reflect variation in risk factors for COVID-19 hospitalisation and disease severity, which would affect the proportion of patients in CC (38). While we can indeed get a better fit by adjusting this (Supplementary Figure 5), leading to a range of values (8.9%-21.0%, mean 14.2%) which are consistent with our meta-analysis on proportion of patients going to CC (mean 14.01%, s.d. 7.73%) (Supplementary Table 5), this is overall not as successful compared to adjusting LoS (Supplementary Table 4). These proportions are likely to be strongly age-dependent, but variation in age distribution between Regions (except London) are unlikely to be substantial enough to drive the observed differences. On the other hand, hospital resource availability does vary by Region: COVID-19 case prevalence alongside case management and bed availability may have affected bed pathways and hence LoS. For context, there were approximately 5,900 critical care beds available in England as of February 2020 (4), and 92,500 ward beds as of June 2020 (39), but these were not evenly distributed between Regions, nor was the COVID-19 case burden (40).

At the NHS Trust level, estimated differences in LoS are harder to interpret. In a single region, Trusts may form a network where each Trust fulfills a specific role, such as specialising in the care of critically ill patients, leading to patient flows between hospitals (41). This could cause the proportion of patients in CC to vary heavily between Trusts, whilst for simplicity this value is kept constant during our fitting process. Therefore, our estimated LoS for each Trust obtained by fitting should not be considered as truly representative. Instead, they serve to show that there is likely substantial variation not only between NHS Regions, but also within. The exact nature of this heterogeneity must then be clarified directly using Trust data on patients LoS.

A greater issue is likely to be inconsistency between data definitions (such as “critical care”) and sources of data (e.g. which Trusts are included) (29). In the bed occupancy dataset, CC beds are defined as “beds which are capable of delivering mechanical ventilation” (29). This may be more restrictive, as beds delivering mechanical ventilation would be ICU beds only (42), while the CO-CIN definition of CC beds includes both ICU and HDU beds (13). However, as the definition uses the word “capable”, it could include HDU beds. Altering the definition of CC beds to account for them being potentially too restrictive would not improve our national model estimates, as it would only further increase the model-predicted ward occupancy, which is already over-estimated.

### 5. Next steps

Bed occupancy predictions crucially rely on predictions of hospital admissions, which are difficult to produce, especially at lower levels, such as the hospital. Using local past estimates of LoS can help narrow down uncertainty, though a key driver will be admission prediction variability. For example, the timing of the UCH & England peak bed occupancy were quite different. Our model would rely on good bed admission predictions to forecast bed occupancy alongside accurate LoS values. Without these, errors in timing of peak can result in long tails of errors in levels of bed occupancy.

Understanding the reasons for the regional and sub-regional heterogeneity is a key next step. Matching this to known risk factor variation (e.g. in socio-economic status and comorbidities) could improve our understanding of clinical care impact. Future work clarifying the structure of patient bed pathways could be used to better understand the factors that contribute to patient outcomes, such as whether a patient is likely to die or be discharged at the end of their hospitalisation (43).

Our model could be expanded to consider additional resource complexity (such as staffing needs) and spare capacity requirements alongside exploring trigger times for the need of future beds could be explored with simple adaptations to this model. Similarly, an exploration of time varying elements, such as admission & discharge procedures, alongside COVID-19 prevalence would be important.

This model structure is not linked specifically to the English situation and hence could be adapted to any setting. We found that patient bed pathways are likely to be similar globally but further data is required on length of stay to parameterise the bed pathways.

### 6. Conclusion

Our results emphasise the importance of local knowledge in predicting bed occupancy and hence capacity planning. We found that using only average LoS by bed types could underestimate bed occupancy at the hospital level. There may be important underlying heterogeneities in LoS, which should be further investigated as they may provide insight into the prevalence of risk factors for COVID-19 or clinical care disparities. As cases of COVID-19 are currently increasing again in England, and for future epidemic preparedness, it is essential to develop the best possible understanding of the bed pathways of patients with COVID-19 now, to avoid local under- or overestimates of forecasted bed occupancy.

## Data Availability

This work uses data provided by patients and collected by the NHS as part of their care and support #DataSavesLives. The CO-CIN data was collated by ISARIC4C Investigators. ISARIC4C welcomes applications for data and material access through our Independent Data and Material Access Committee (https://isaric4c.net).
The code for the bed occupancy model and bed pathways data are available in a GitHub repository (https://github.com/qleclerc/COVID_bed_occupancy).

https://github.com/qleclerc/COVID_bed_occupancy

## Acknowledgements

We would like to thank University College Hospitals NHS Foundation Trust for their provision of data. Initially this was made available to us to support UCH bed occupancy prediction modelling in the early COVID-19 phase of 2020.

This work uses COCIN data provided by patients and collected by the NHS as part of their care and support #DataSavesLives. We are extremely grateful to the 2,648 frontline NHS clinical and research staff and volunteer medical students, who collected this data in challenging circumstances; and the generosity of the participants and their families for their individual contributions in these difficult times. We also acknowledge the support of Jeremy J Farrar and Nahoko Shindo.

## Copyright

This is an open access article distributed in accordance with the terms of the Creative Commons Attribution (CC BY 4.0) license, which permits others to distribute, remix, adapt and build upon this work, for commercial use, provided the original work is properly cited. See: http://creativecommons.org/licenses/by/4.0.

The Corresponding Author has the right to grant on behalf of all authors and does grant on behalf of all authors, an exclusive licence (or non exclusive for government employees) on a worldwide basis to the BMJ Publishing Group Ltd to permit this article (if accepted) to be published in BMJ editions and any other BMJPGL products and sublicences such use and exploit all subsidiary rights, as set out in our licence.

## Transparency declaration

The lead author affirms that this manuscript is an honest, accurate, and transparent account of the study being reported; that no important aspects of the study have been omitted; and that any discrepancies from the study as planned (and, if relevant, registered) have been explained.

## Ethical approval

Ethical approval for ISARIC was given by the South Central - Oxford C Research Ethics Committee in England (Ref 13/SC/0149), the Scotland A Research Ethics Committee (Ref 20/SS/0028) and the WHO Ethics Review Committee (RPC571 and RPC572, 25 April 2013).

Local ethical approval for the use of the UCH data was provided by the London School of Hygiene and Tropical Medicine Ethics Committee (Ref 21907) for data analysis to provide a service evaluation.

## Registration

The ISARIC WHO CCP-UK study was registered at https://www.isrctn.com/ISRCTN66726260 and designated an Urgent Public Health Research Study by NIHR.

## Data availability

This work uses data provided by patients and collected by the NHS as part of their care and support #DataSavesLives. The CO-CIN data was collated by ISARIC4C Investigators.

ISARIC4C welcomes applications for data and material access through our Independent Data and Material Access Committee (https://isaric4c.net).

The code for the bed occupancy model and bed pathways data are available in a GitHub repository (https://github.com/qleclerc/COVID_bed_occupancy).

## Patient and Public Involvement

Patients or the public were not involved in the design, or conduct, or reporting of this study. As such, dissemination of the results to these groups is not applicable.

