## Supplementary Material for "Importance of patient bed pathways and length of stay differences in predicting COVID-19 bed occupancy in England"

### Review of percentage of patients with COVID-19 that need ICU care

In order to obtain an international perspective on the proportion of patients entering each bed pathway we used a systematic review of length of stay for patients with COVID-19 as a dataset for studies likely to contain this information. The references in this systematic review (1) were gathered up to 12th April 2020 and thus cover the first period of the pandemic.

Each of the references cited in the paper were screened for information on the total number of patients included in the study, and how many of these were admitted to a critical care bed. In total, 21 relevant papers were found. None reported bed pathway proportions, only proportion entering a critical care bed. All of these studies only used the term “intensive care unit” (ICU) to refer to critical care, and did not mention high dependency units.

The studies found cover several countries from late December 2019 to the start of April 2020 with the number of patients per study varying from 28 to 3316. Due to this wide range, a weighted mean and weighted standard deviation was calculated for the percentage of patients transferred to an ICU (Supplementary Table 1). However, the majority of these studies only cover China (13/21), which may bias our results. All calculations were performed in R (2).

**Supplementary Table 1: Percentage of patients with COVID-19 directed to ICU care from 21 studies.** Mean and standard deviation are weighted by the number of patients in each study.

| **Reference** | **Country** | **Number of patients in study** | **Percentage to ICU** |
| --- | --- | --- | --- |
| Liu et al (3) | China | 64 | 0 |
| Ludvigsson, J (4) | China | 171 | 2 |
| Petrilli et al (5) | USA | 2741 | 4 |
| Guan et al (6) | China | 1099 | 5 |
| Chen et al (7) | China | 249 | 9 |
| Cai et al (8) | China | 298 | 10 |
| Zhang et al (9) | China | 28 | 11 |
| Inciardi et al (10) | Italy | 99 | 12 |
| Richardson et al (11) | USA | 2634 | 14 |
| Cao et al (12) | China | 102 | 18 |
| ISARIC Report (13) | Various | 3316 | 20 |
| Qi et al (14) | China | 267 | 20 |
| Zhang et al (15) | China | 221 | 20 |
| Rodriguez-Morales et al (16) | Various | 656 | 20 |
| Lewnard et al (17) | USA | 617 | 26 |
| Zhou et al (18) | China | 191 | 26 |
| Wu et al (19) | China | 201 | 26 |
| Wu et al (20) | China | 188 | 27 |
| Zaninotto et al (21) | Italy | 75 | 28 |
| Chao et al (22) | USA | 46 | 28 |
| Huang et al (23) | China | 41 | 32 |
| **Weighted Mean** |  |  | *14.01* |
| **Weighted Standard Deviation** |  |  | *7.73* |

### Sensitivity analysis

**Supplementary Table 2: Patient bed pathways and length of stay for patients with COVID-19 from University College Hospital (UCH) and the COVID-19 Clinical Information Network (CO-CIN).** CC: critical care. n: number of occurrences of that pathway (for Bed pathways), or bed type (for Averages). Note that the sum of n for the bed pathways differs from the sum for the averages, since two stages of the same bed type in one pathway correspond to two occurrences of that bed type in the averages. IQR.: interquartile range.

| **Dataset** |  | **Beds** | **n** | **Proportion** | **Stage 1** | | **Stage 2** | | **Stage 3** | | **Total** | |
| --- | --- | --- | --- | --- | --- | --- | --- | --- | --- | --- | --- | --- |
|  |  |  |  |  | **Median** | **IQR** | **Median** | **IQR** | **Median** | **IQR** | **Median** | **IQR** |
| **UCH** | **Bed pathways** | CC | 8 | 0.048 | 4.88 | 3.36 - 6.12 | / | / | / | / | 4.88 | 3.36 - 6.12 |
|  |  | CC, Ward | 4 | 0.024 | 3.71 | 2.27 - 5.78 | 2.38 | 1.74 - 3.79 | / | / | 8.46 | 6.78 - 9.18 |
|  |  | Ward | 137 | 0.815 | 3.36 | 1.93- 5.53 | / | / | / | / | 3.36 | 1.93- 5.53 |
|  |  | Ward, CC | 9 | 0.053 | 1.23 | 1.10 - 1.49 | 5.11 | 3.08 - 5.77 | / | / | 5.94 | 5.52 - 8.18 |
|  |  | Ward, CC, Ward | 10 | 0.060 | 1.95 | 0.82 - 3.24 | 3.55 | 3.04 - 4.40 | 2.12 | 2.01 - 2.78 | 7.92 | 6.85 - 8.44 |
|  | **Averages by bed type** | CC | 31 | 0.154 | 4.05 | 2.83 - 5.61 | / | / | / | / | 4.05 | 2.83 - 5.61 |
|  |  | Ward | 170 | 0.846 | 2.91 | 1.64 - 5.23 | / | / | / | / | 2.91 | 1.64 - 5.23 |
|  | **Total** | All | 168 | 1 | / | / | / | / | / | / | 3.97 | 2.10 - 6.85 |
| **CO-CIN** | **Bed pathways** | CC | 232 | 0.006 | 7 | 4 - 14 | / | / | / | / | 7 | 4 - 14 |
|  |  | CC, Ward | 2,521 | 0.069 | 9 | 4 - 19 | 3 | 0.25 - 9 | / | / | 16 | 8.25 - 28 |
|  |  | Ward | 29,975 | 0.821 | 6 | 3 - 12 | / | / | / | / | 6 | 3 - 12 |
|  |  | Ward, CC | 183 | 0.005 | 2 | 1 - 4 | 5 | 2 - 10 | / | / | 8 | 4 - 14 |
|  |  | Ward, CC, Ward | 3,603 | 0.099 | 2 | 1 - 4 | 8 | 4 - 17 | 3 | 0.25 - 8 | 18.25 | 10.25 - 31 |
|  | **Averages by bed type** | CC | 6,539 | 0.141 | 8 | 4 - 17 | / | / | / | / | 8 | 4 - 17 |
|  |  | Ward | 39,885 | 0.859 | 5 | 2 - 11 | / | / | / | / | 5 | 2 - 11 |
|  | **Total** | All | 36,514 | 1 | / | / | / | / | / | / | 7.25 | 3 - 15 |

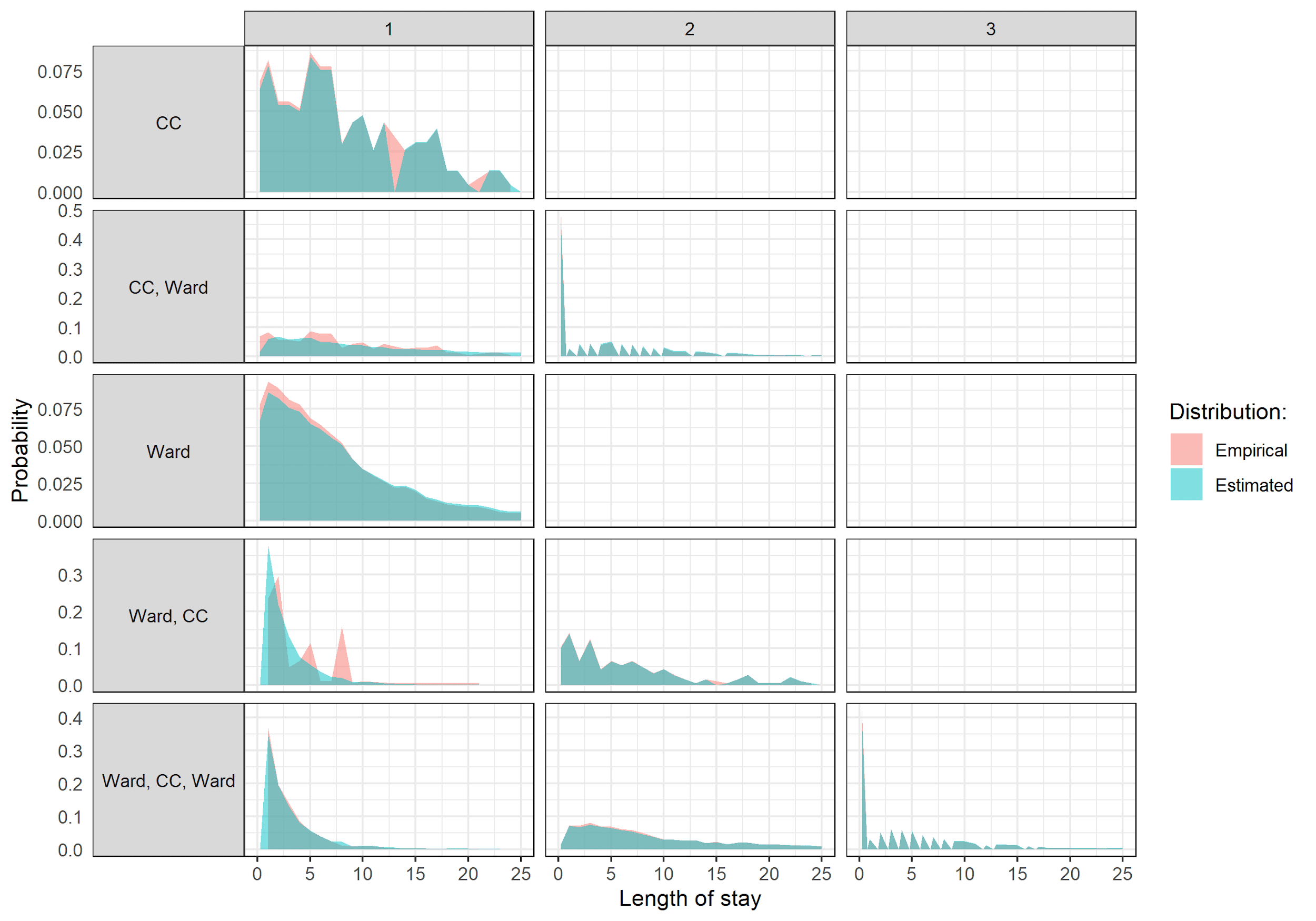

**Supplementary Figure 1: Empirical and estimated distributions for the length of stay values in CO-CIN patient bed pathways.** Estimated distributions were obtained by multi-state modelling, to correct for patients without complete follow-up. Distributions are presented by bed pathways (rows) and stages in the pathway (columns).

**Supplementary Table 3:** **Squared difference between model-predicted UCH bed occupancy and UCH data, under different assumptions on LoS rounding, LoS distribution, and using bed pathways instead of averages by bed type.** Results are from 100 model runs.

| **LoS rounding** | **Distribution** | **LoS** | **Squared difference** |
| --- | --- | --- | --- |
| Round | Weibull | Average LoS | 2667.14 |
|  |  | Bed pathways | 916.03 |
|  | Lognormal | Average LoS | 3665.66 |
|  |  | Bed pathways | 5698.71 |
| Ceiling | Weibull | Average LoS | 1431.08 |
|  |  | Bed pathways | 586.77 |
|  | Lognormal | Average LoS | 3888.02 |
|  |  | Bed pathways | 7311.79 |

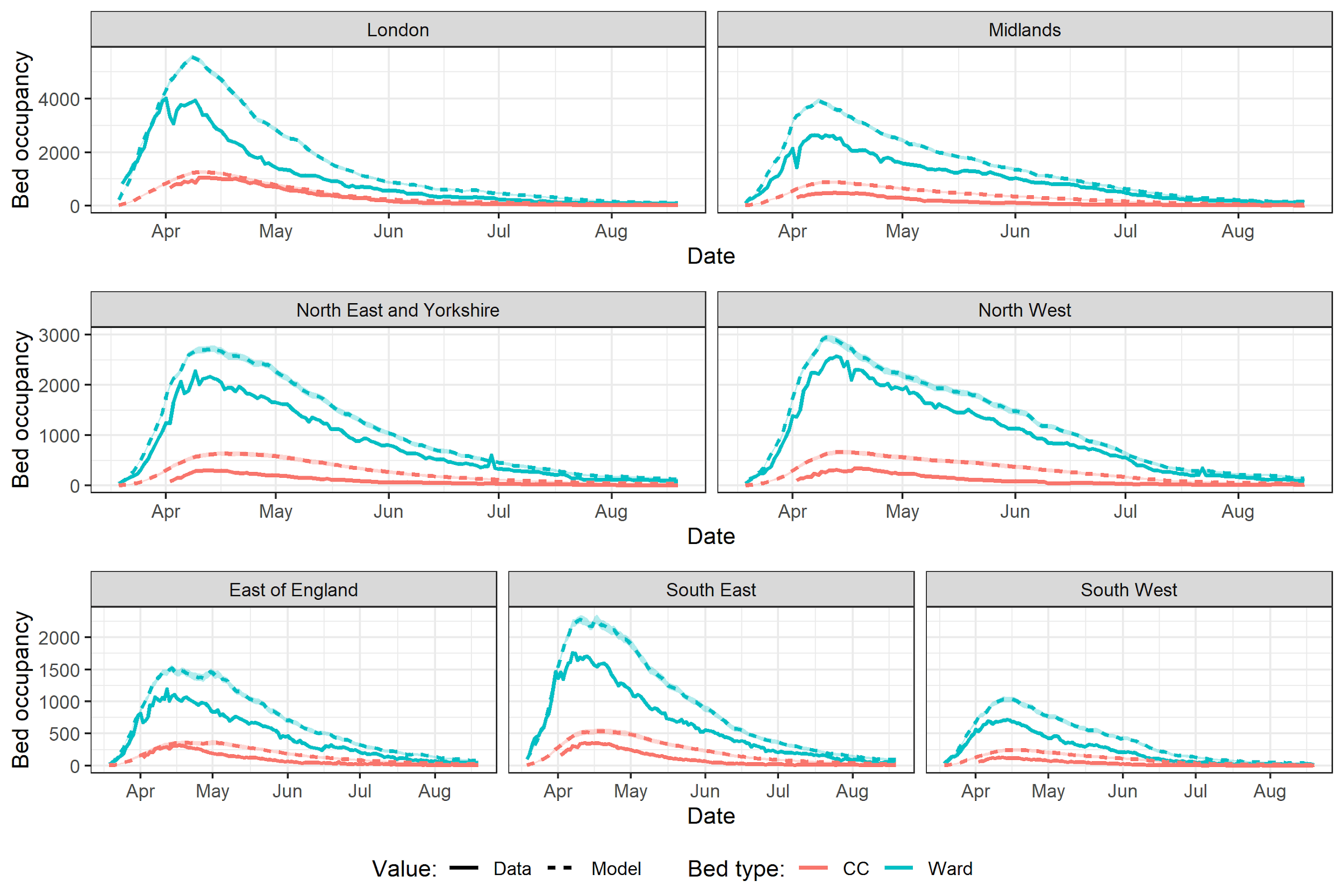

**Supplementary Figure 2: Model-predicted bed occupancy at the NHS Region level using bed pathways LoS values from CO-CIN.** Dotted line shows true bed occupancy for the same period according to publicly available hospitalisation data. CC: critical care. Results are from 100 model runs.

**Supplementary Table 4:** **Squared difference (x10^5) between model-predicted bed occupancy and NHS Regions data, using CO-CIN LoS values, best-fit LoS values, and best-fit average proportion of patients staying in a CC bed.** Results are from 100 model runs.

| **NHS Region** | **CO-CIN average LoS** | **CO-CIN bed pathways LoS** | **Best-fit average LoS** | **Best-fit average proportion to CC** |
| --- | --- | --- | --- | --- |
| East of England | 22.00 | 62.50 | 6.42 | 6.99 |
| London | 262.51 | 553.09 | 48.90 | 67.23 |
| Midlands | 173.03 | 356.06 | 69.98 | 81.66 |
| North East and Yorkshire | 85.29 | 207.31 | 17.86 | 23.25 |
| North West | 56.58 | 142.66 | 21.32 | 17.60 |
| South East | 47.08 | 125.04 | 12.76 | 13.95 |
| South West | 11.16 | 28.10 | 4.46 | 4.16 |

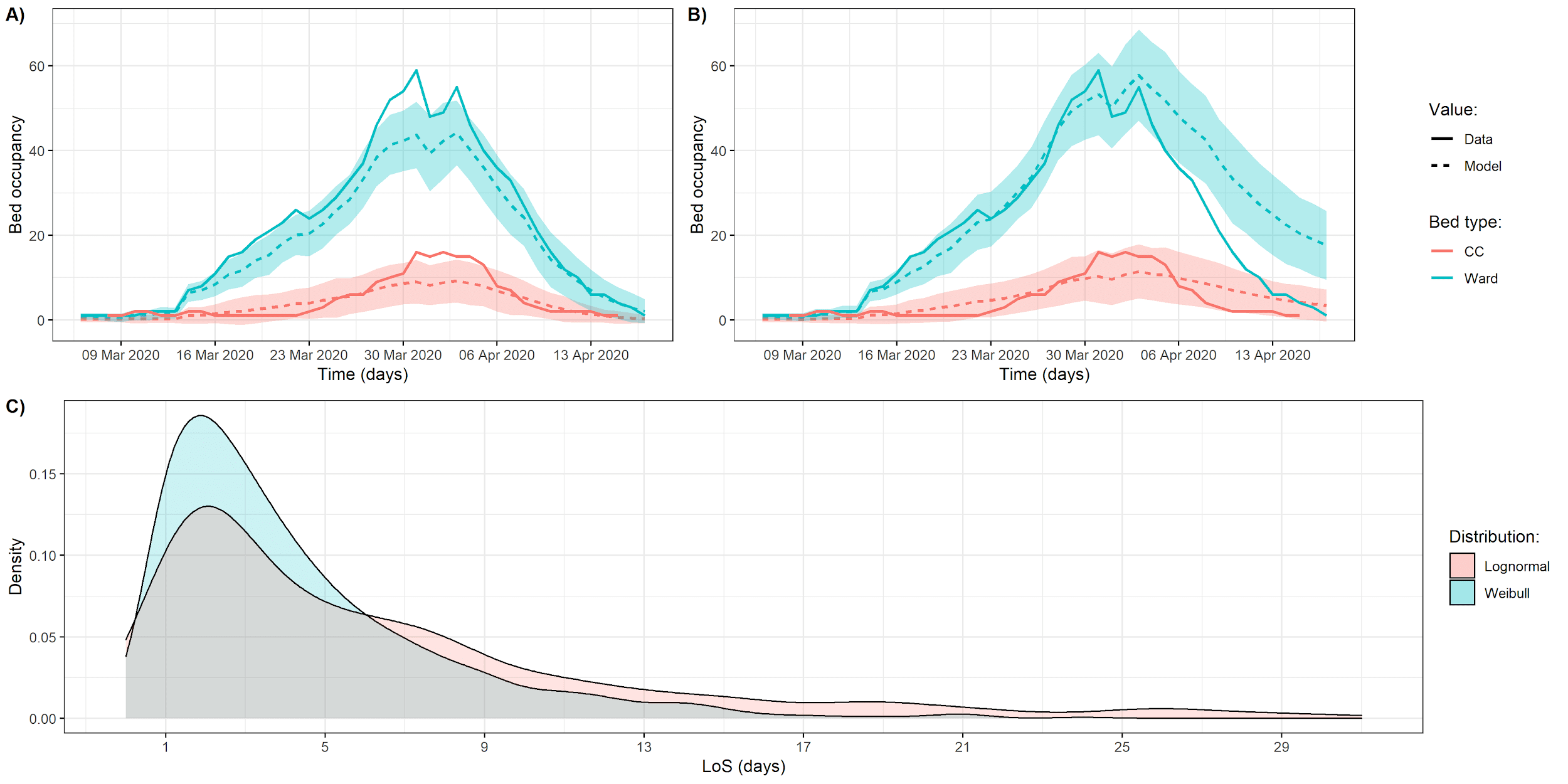

**Supplementary Figure 3: UCH bed occupancy predictions are more accurate when using a Weibull instead of a Lognormal distribution for length of stay.** **A)** Bed occupancy at UCH and model-predicted bed occupancy using UCH average length of stay estimates and a Weibull distribution. **B)** Bed occupancy at UCH and model-predicted bed occupancy using UCH average length of stay estimates and a Lognormal distribution. **C)** Weibull and Lognormal distributions for ward bed LoS, using UCH parameters (mean: 3.90, standard deviation: 3.68). Results are from 100 model runs.

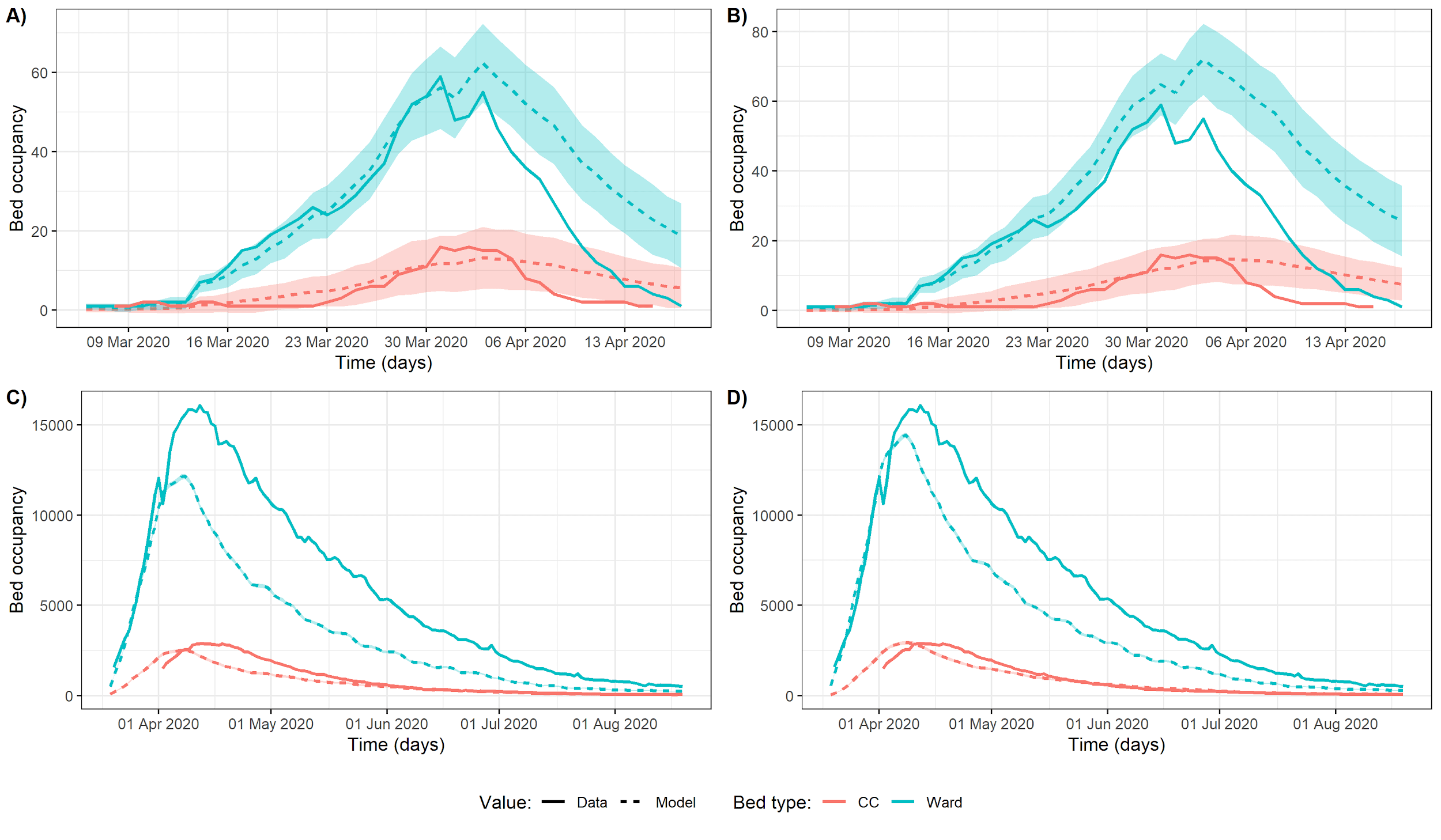

**Supplementary Figure 4: Using UCH LoS estimates to predict England bed occupancy, or CO-CIN LoS estimates to predict UCH bed occupancy, respectively lead to an underestimate and an overestimate of true bed occupancy.** Bed occupancy at UCH and model-predicted bed occupancy using CO-CIN **A)** average length of stay estimates or **B)** bed pathways. Bed occupancy in England and model-predicted bed occupancy using UCH **C)** average length of stay estimates or **D)** bed pathways. Shaded area is the 95% confidence interval from 100 model runs. Note that the time period is different between data from UCH and England.

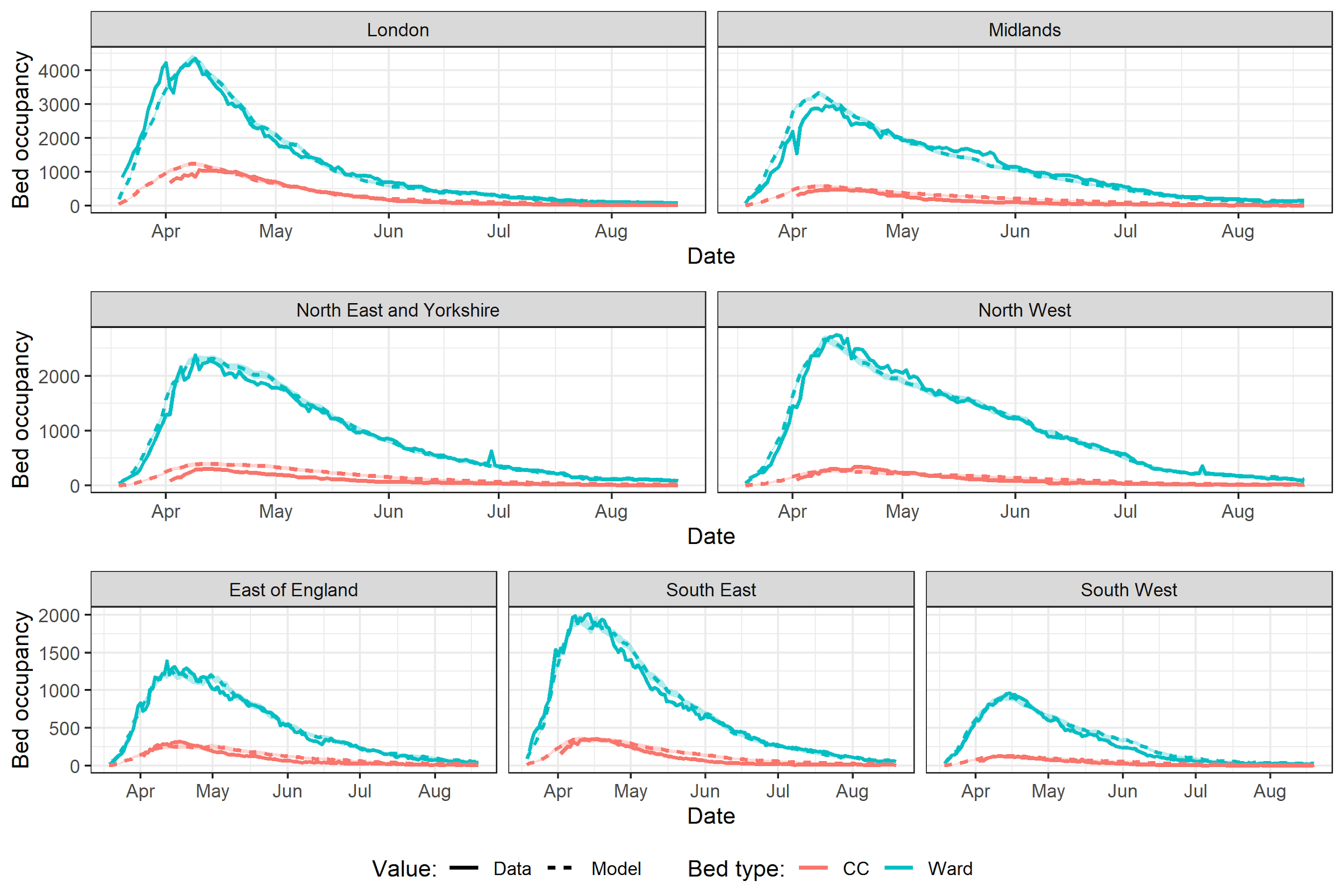

**Supplementary Figure 5: Model-predicted bed occupancy at the NHS Region level using best-fit proportion of patients requiring a critical care (CC) bed.** Best-fit proportion to CC values were obtained by minimising the sum of squared differences between model values and data, for each Region separately. Results are from 100 model runs. Best-fit proportions are provided in Supplementary Table 3.

**Supplementary Table 5: Best fitting proportion of patients requiring either a ward or critical care (CC) bed by NHS Regions.** The England weighted average is the average of fitted proportions to ward or CC in NHS Regions, weighted by the proportion of cumulative England hospitalisations that occured in each Region. The length of stay values are fixed to the CO-CIN averages by bed type.

| **Fitting type** | **Geography** | **Proportion to ward** | **Proportion to CC** |
| --- | --- | --- | --- |
| **CO-CIN values** | *England average* | *0.856* | *0.144* |
| **Best fit by Region** | East of England | 0.841 | 0.159 |
|  | London | 0.790 | 0.210 |
|  | Midlands | 0.858 | 0.142 |
|  | North East and Yorkshire | 0.864 | 0.136 |
|  | North West | 0.911 | 0.089 |
|  | South East | 0.858 | 0.142 |
|  | South West | 0.887 | 0.113 |
|  | *England weighted average* | *0.853* | *0.147* |
